# Reimagining Cryptogenic Stroke Care: Collaborative Care and Inpatient Insertable Cardiac Monitors for Detection of Atrial Fibrillation

**DOI:** 10.1101/2024.08.15.24312089

**Authors:** Nabeel A. Herial, Daniel R. Frisch, Elan Miller, Priyadarshee Patel, Alfredo Munoz, Melissa Warren, Jane Khalife, Shyam Majmundar, Nathan Farkas, Shaista Alam, Robin Dharia, Diana Tzeng, Behzad B. Pavri, Reginald T. Ho, Arnold Greenspon, Rodney Bell, Pascal Jabbour, Robert Rosenwasser

## Abstract

**Background:** Atrial fibrillation (AF) is a known risk factor of ischemic stroke and AF-related stroke is twice more likely to be fatal. Long-term cardiac rhythm monitoring using insertable cardiac monitors (ICMs) has greater diagnostic yield compared to conventional monitoring in detecting AF, and clinical utility of ICMs is established in cryptogenic stroke, strokes due to large artery atherosclerosis and small vessel disease. A registry-based study was conducted to evaluate inpatient implantation of ICMs and feasibility of vascular and interventional neurologists as implanters using novel collaborative clinical care pathway for cryptogenic stroke.

**Methods:** Multiyear data from a hospital-based registry at a comprehensive stroke center was reviewed to evaluate inpatient ICM implantation and test feasibility of vascular and interventional neurologists/VIN as implanters of ICMs together with cardiology using a novel collaborative care pathway. Reviewed data included number of ICMs, implantation trend, inpatient vs. outpatient setting, time to ICM implantation, inpatient workflow including defined roles of team members, and AF detection rate.

**Results:** Total of 290 ICMs for cryptogenic stroke were implanted when patients where in the hospital and 78 as outpatient after discharge during the study period of 3 years. The majority of inpatient ICM implants were performed by VIN (n=181) and ICM use for cryptogenic stroke increased by 130%. Average time to inpatient ICM implant was 4.1 days with 77% in 5 and 95.5% within 10 days post-stroke. Average time to out-patient ICM placement was 57 days. AF detection rate of 36.5% was noted at 24 months with a collaborative care pathway.

**Discussion:** Inpatient implantation of ICMs is feasible and was performed safely and efficiently by VIN together with cardiology using a collaborative care pathway. Increase in utilization of ICMs and higher AF detection rates were noted. Findings support innovative efforts to improve access and close the gaps in delivery of cryptogenic stroke care to ultimately reduce the secondary stroke burden.

**Clinical Perspective:**

- Direct inpatient implantation of ICMs in cryptogenic stroke allows increased rates of early long-term cardiac rhythm monitoring and detection of AF.
- Collaborative clinical practice with inpatient ICM implantation by vascular and interventional neurologists is feasible and safe. This novel approach could potentially increase access to cardiac rhythm monitoring and exemplify patient-centered care.

## Introduction

Stroke remains a leading cause of death worldwide and incidence of ischemic stroke which accounts for most of the disease burden, is projected to increase to 9.6 million in 2030. ^1, 2^ The frequently documented etiologies of ischemic stroke include large-artery atherosclerosis, small-vessel disease, and cardioembolism, ^3^ and about one-fourth of ischemic strokes are referred to as cryptogenic where diagnostic evaluations reveal no probable cause of stroke. ^4^ Atrial fibrillation (AF) is a well-known risk factor of ischemic stroke, ^5, 6^ and evaluation for subclinical AF in cryptogenic stroke with long-term cardiac rhythm monitoring is recommended. ^7^ Recent evidence revealed AF was also more common in ischemic strokes attributed to large- or small-vessel disease. ^8^

Detection of subclinical AF is essential in the secondary prevention of ischemic stroke with timely initiation of treatment with anticoagulation. In cryptogenic stroke patients with no contraindications for anticoagulation, the AHA/ASA guidelines recommend long-term cardiac rhythm monitoring for detection of AF, ^9^ and evidence suggests use of Insertable Cardiac Monitors (ICMs) for long-term cardiac rhythm monitoring provides the highest diagnostic yield in cryptogenic stroke. ^7^ The 2022 European Stroke Organization (ESO) guidelines and the consensus statement recommends use of ICMs compared to non-implantable external monitoring devices to increase detection of AF in cryptogenic stroke. ^10^

Evaluation of cryptogenic stroke patients with long-term cardiac rhythm monitoring involves coordinated care between multiple clinical specialties. In this study, we evaluated inpatient implantation of ICMs and role of vascular and interventional neurologists as implanters using a novel collaborative clinical care pathway for cryptogenic stroke.

## Methods

The study was conducted at a comprehensive stroke center of a university health system after approval from the Institutional Review Board. This is an observational study of data obtained from a hospital registry of ICM implants performed for evaluation of cryptogenic stroke patients. STrengthening the Reporting of OBservational studies in Epidemiology (STROBE) guidelines were used in outlining the methods and findings of the study. Data collected includes patient identifiers and sensitive device implant information and can be made available from the corresponding author upon a reasonable request. A novel clinical care pathway was developed and implemented at the hospital for Neuroscience, a facility reserved for acute care of Neurological and Neurosurgical patients. The pathway was developed with collaboration between clinical specialties of cardiac electrophysiology (EP), vascular and interventional neurology (VIN) and neurosurgery (NS) for implantation of ICMs (Figure 1).

**Figure 1:**
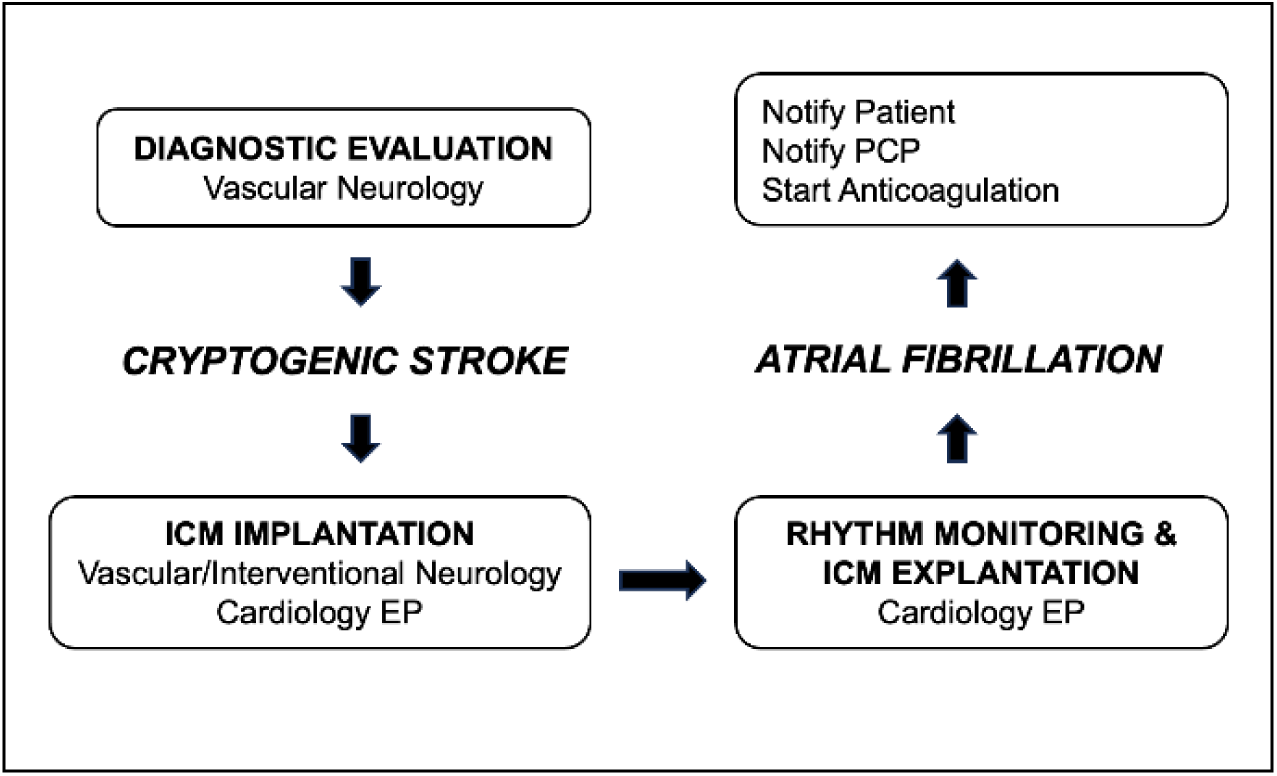
Collaborative Clinical Care Pathway

Primary objectives of the study were to facilitate evaluation of cryptogenic stroke patients by early initiation of long-term cardiac rhythm monitoring with direct inpatient implantation of ICMs prior to discharge using a clinical care pathway, test the feasibility of a novel practice model with VIN as implanters of ICMs, and implement a framework for inpatient and outpatient care and communication.^11^ The secondary objectives included evaluating the safety and complication rate of inpatient ICM implantation and AF detection rate in the group. The diagnosis of cryptogenic stroke was determined by the attending physician of the inpatient stroke unit. A total of five vascular neurologists with subspecialty training and board-certification were involved in the evaluation and diagnosis of cryptogenic stroke in patients admitted to the stroke unit of the Neuroscience hospital. Continuous cardiac rhythm monitoring via telemetry was performed as standard evaluation in all stroke patients during hospitalization. For eligible cryptogenic stroke patients with no contraindications for anticoagulation, an electronic and direct verbal referral is sent to an EP or VIN for implantation of ICM. A preferred age cut-off of ≥60 years was considered for implant and ICM implantation was offered regardless of the patient’s insurances status. Seven eligible inpatients (2%) refused ICM implant, stating personal preference as the reason for opting an external device over a subcutaneous device implant for cardiac rhythm monitoring. In the inpatient setting, VIN physician with privileges approved by the hospital credentialing committee performed the procedure. VIN physicians were required to meet all requirements including performing a required minimum number of 20 ICM implant procedures under supervision of a EP physician.

After determination of ICM as medically reasonable and necessary for further evaluation of the stroke, an informed consent from the patient and/or family was obtained. ICM implants were preferentially performed by VIN on the day of discharge from the hospital. Implantation and clinical indication-based programing of device was performed by VIN. Registration of device-related information with cardiology EP section was simultaneously performed to facilitate outpatient follow up and continuity of care. Regardless of the clinical specialty implanting the ICM, all inpatients after discharge were scheduled to follow up in the EP clinic and interpretation of ICM data and ECG tracings was performed by EP section. Inpatients who were determined eligible for long-term cardiac rhythm monitoring but did not receive an ICM, were discharged with Mobile Cardiac Outpatient Telemetry (MCOT) for external monitoring for 21-days with outpatient evaluation in the EP clinic. All outpatient ICM implants were exclusively performed by EP. Long-term monitoring of the cardiac rhythm data from all ICM devices was done by the EP section and a group notification of AF alert was triggered when data a review revealed evidence of AF (Figure 2). Patients and their primary care physicians were notified of the new diagnosis and anticoagulation was initiated immediately for secondary prevention of ischemic stroke if there were no contraindications.

**Figure 2:**
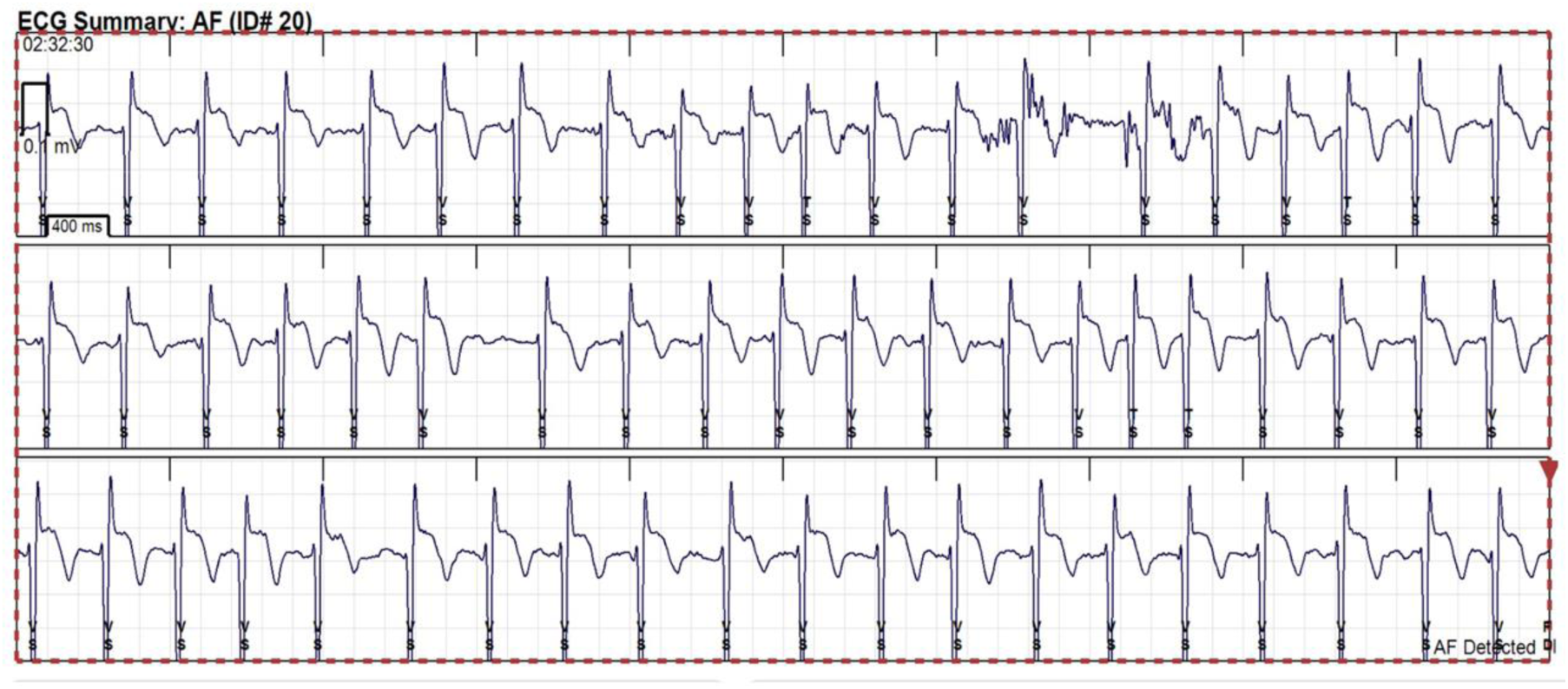
Illustration of an episode of atrial fibrillation characterized by irregularly irregular rhythm and absence of a P wave as detected on an implanted insertable cardiac monitor.

### ICM implantation procedure

After informed consent was obtained from the patient and/or family, procedure was performed under sterile conditions using local anesthesia. With patient in supine position, the precordial area was prepped, and left 4^th^ intercostal space was localized by palpation. At approximately 2 cm lateral to the left sternal border, using a sterile hypodermic needle, skin was infiltrated with 10-20 cc of local anesthetic (1% lidocaine). A small incision of ∼1 cm was made using a supplied angled blade in the plane parallel to the chest wall. The ICM device was inserted within the subcutaneous space, at an angle of ∼45 degrees to the sternum. Device was then programmed for detection of atrial tachycardias, including fibrillation using cryptogenic stroke as the clinical indication.

Amplitude and sensitivity of the waveform was evaluated and after confirming an optimal signal (preferably >20 mV/µV) from the device, the incision was secured with adhesive bandage (Steri-Strips) or absorbable sutures. In uncommon instances of a suboptimal device signal, removal and repositioning of the device was done. Complications associated with the procedure such as hematoma, infection/abscess, device rejection phenomenon with or without tissue reaction, skin erosion, and device migration were recorded.

A multidisciplinary team was involved in implementing the pathway and roles of all members was clearly outlined. ICM implant procedures were mostly performed in the controlled-access, peri-procedural sterile areas of the interventional neuroradiology suite with assistance from the interventional staff and nurse practitioners. In-service education on ICMs implantation procedure was offered to the stroke unit and interventional neuroradiology nursing staff. Education of the staff at the acute rehabilitation centers receiving the discharged stroke patients was also performed.

For the billing of the procedure, CPT^®^ code 33285 for ‘Insertion, subcutaneous cardiac rhythm monitor, including programming’ and ICD-10-PCS code 0JH632Z for ‘Insertion of monitoring device into chest subcutaneous tissue and fascia, percutaneous approach’ were used. Reimbursement by Medicare for inpatient hospital services is based on a classification system known as Medicare severity diagnosis related groups (MS-DRG) and a single payment is made for all procedures and supplies related to an inpatient stay and the assigned MS-DRG (https://www.cms.gov/icd10m/version39-fullcode-cms/fullcode_cms/P0001.html). Frequently assigned MS-DRG codes for implantable loop recorders included 040, 041, and 042 based on presence or absence of major complication and comorbidity (MCC) or complication and comorbidity (CC). Each MS-DRG has a relative weight that is converted to a flat payment to the facility and a single MS-DRG is assigned for each inpatient stay irrespective of the number of procedures performed. For patients with no qualifying inpatient procedure, ICM implant allows hospitals to use a procedural MS-DRG for the services provided. Physician reimbursement is based on the CPT codes for the procedure, irrespective of the setting where service is provided: inpatient, outpatient or the office. According to the Medicare Physician Fee Schedule (MPFS) of 2024, CPT code 33285 is assigned total relative value units (RVUs) of 2.57 for the physician payment.

### Data Analysis

Three-year data from the hospital registry of ICM implantation was reviewed (January 1, 2017 to December 31, 2019). A total of 2,744 ischemic stroke patients were discharged from the Neuroscience hospital of the health system during the study period of 3 years. Based on a conservative estimate of cryptogenic stroke prevalence of 15% of all ischemic strokes, a minimum of 75 ICM implants/year were estimated. Collaborative care pathway with VIN as implanters of ICM was implemented in the 4^th^ quarter of 2017. To account for any potential variability in the stroke admissions and to examine the trends, quarterly data from the registry before and after implementation of the clinical pathway was compared. Total number of ICM implants, procedures performed by VIN and EP, complication rates, AF detection rate, patient characteristics including demographics, echocardiographic features were examined. Numerical and categorical data were presented as mean ± SD and frequencies, respectively. Comparison between groups was performed using independent sample t-test or Wilcoxon rank sum test, or Chi-square (χ2) test, based on the data distribution. Statistical hypotheses were tested at 0.05 level of significance (α) and analyses performed using SPSS 24.0.

## Results

A total of 428 ICMs were implanted during the study period of 3 years. The majority of the ICM implants performed at the neuroscience hospital were for the clinical indication of cryptogenic stroke (n=368, 86%). There were 290 (67.8%) ICMs were implanted in the inpatient setting prior to discharge and 78 (18.2%) were performed on an outpatient basis. Average age of the study patients was 67 years (SD=11) and majority were male (59%). Prevalence of hypertension and diabetes in the study population were 67% and 34% respectively. Current and former smoking was reported in 19.5% and 55% of the patients. Comparison of quarterly data from the registry showed an increase in the number of ICM implants after implementing the collaborative pathway with VIN as ICM implanters. Within the first 3 quarters of implementation, 80 ICM procedures were performed by VIN. Figure 3 shows matched-quarterly data of ICM implants in 2017 by EP and in 2018 by both EP and VIN in collaboration. Nearly, 3-time increase in ICM implants was noted with increased monitoring in each of the matched quarters of 2017 and 2018 (16 vs. 23, p=0.065; 12 vs. 33, p=0.009; and 13 vs. 35, p=0.008).

**Figure 3:**
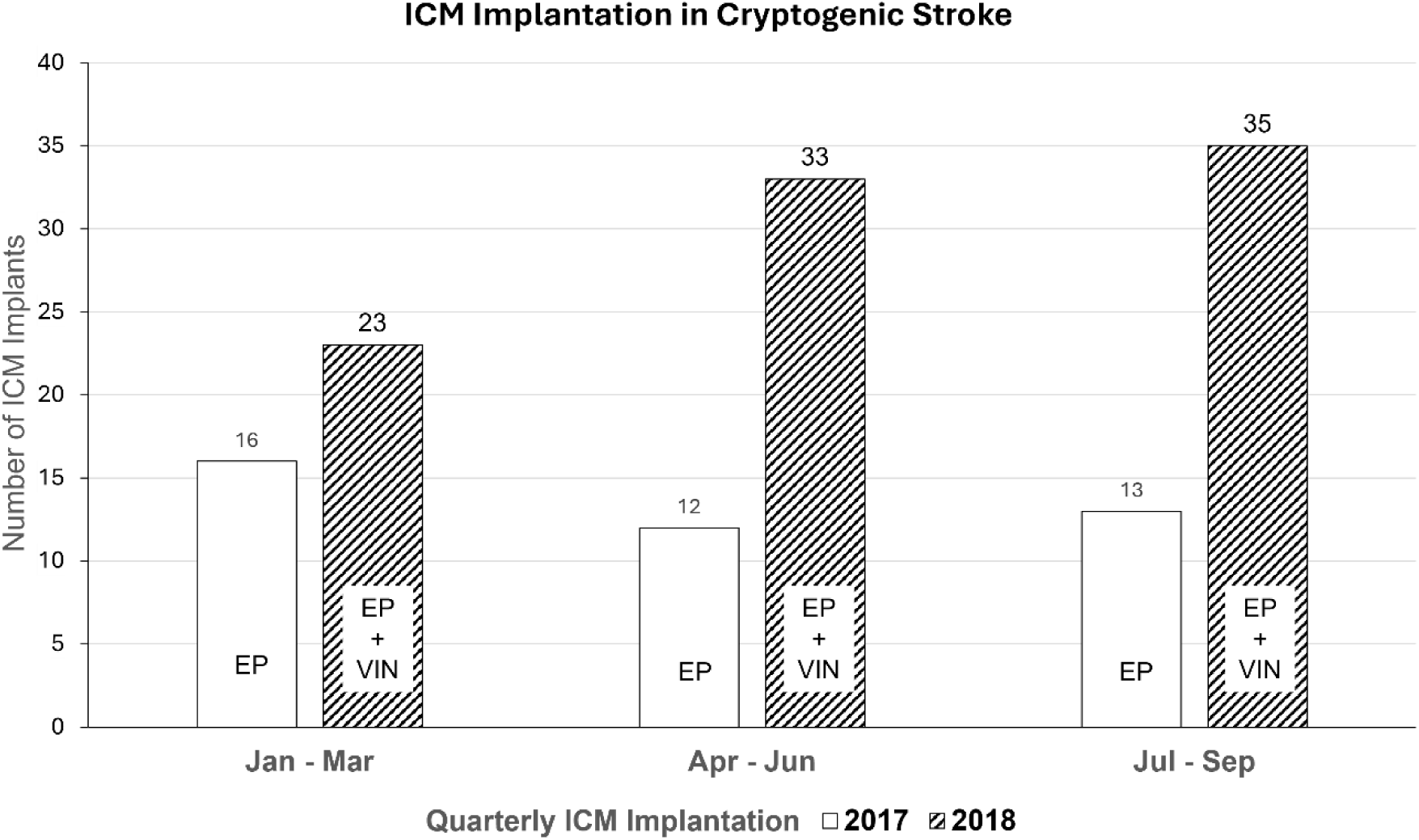
ICM implants before and after implementation of collaborative care pathway for cryptogenic stroke. ICM Insertable Cardiac Monitor, EP Electrophysiology, VIN Vascular and Interventional Neurology.

The average hospital length of stay of the inpatient ICM cohort was 5 days and average time from admission with stroke to ICM implantation was 4.1 days. 77% of ICM implants were done within 5 days, 18.5% in 5-10 days, and <5% in 11 or more days post-admission with stroke. Figure 4 illustrates the frequencies of inpatient and outpatient ICM implants in the study population. Majority of inpatient ICM implants in 2018 and 2019 were performed by VIN, 78% and 80%, respectively.

**Figure 4:**
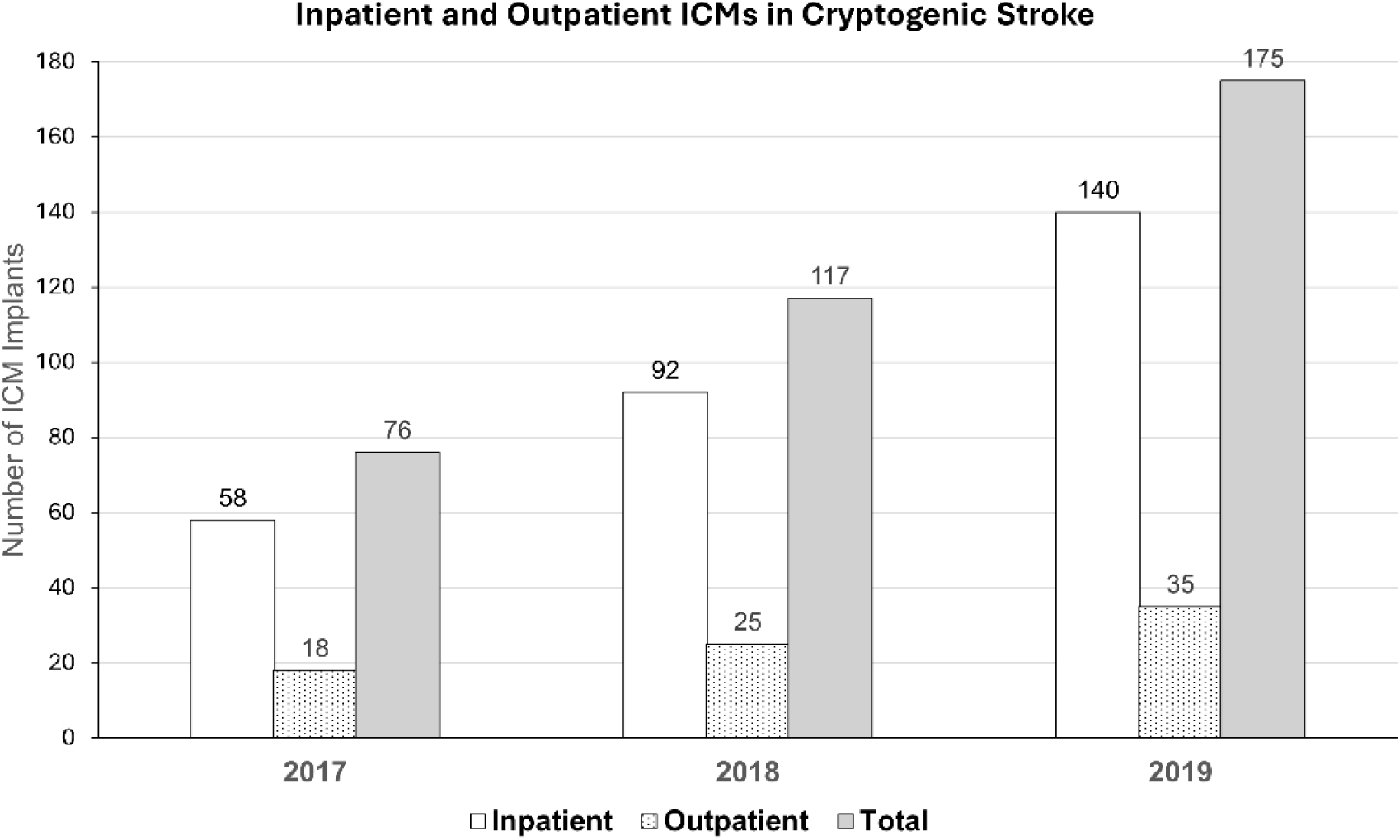
Frequencies of inpatient and outpatient ICM implants

Average time from admission with stroke to outpatient ICM implantation was 57 days. Of the ICMs implanted in the outpatient setting, 16% were done in 15 days, 29% in 30 days, and 53% in more than 30 days after stroke. AF detection rates in the study were 19.6% in 2017, 26% in 2018 and 36.5% in 2019. In the subgroup of cryptogenic stroke patients that underwent ICM implants by VIN as inpatients (in 2018 and 2019), average AF detection rate was 31% and false positive AF alerts was 1.5%. The average time from inpatient ICM implantation to AF detection was 87 ± 43.5 (mean ± sd) days. No procedural complications were reported with inpatient ICM implants performed by VIN.

## Discussion

Detection of subclinical AF has an important clinical implication of change in treatment to anticoagulation for secondary prevention of ischemic stroke. Evidence suggests that in about 88% of patients who had AF, the diagnosis would be missed if the period of electrocardiographic monitoring is limited to 30 days. ^7^ In cryptogenic stroke patients with no contraindications for anticoagulation, long-term cardiac rhythm monitoring for detection of AF is hence recommended by the AHA/ASA. ^9^ Long-term monitoring, specifically with ICMs is proven to have higher diagnostic yield compared to conventional monitoring in the detection of AF. ^12^

A large cohort study evaluating clinical outcomes in cryptogenic stroke showed that initial use of external cardiac monitors was associated with a lower rate of subsequent transition to ICM for long-term monitoring and substantial delays in the diagnosis of AF. ^13^ Proceeding directly to inpatient implantation of ICMs in cryptogenic stroke patients is associated with increased diagnostic efficiency, early diagnosis of AF allowing timely treatment with anticoagulation and better clinical outcomes. ^13, 14^ Furthermore, in a cost comparative analysis, immediate ICM use was associated with cost-savings to both the payers and the patients. ^15, 16^ Inpatient ICM implantation prior to the hospital discharge ensures uninterrupted, high-yield cardiac rhythm monitoring, patient retention in the practice and prevents loss during follow-up. ICM implantation qualifies as a procedure and impacts the MS-DRG for a given hospitalization which results in reimbursement to the hospital and minimizes cost to the patient. The procedure provides an additional opportunity during hospitalization for patient and/or family education.

In this study, we utilized a collaborative care pathway to successfully adopt the approach of inpatient ICM implantation for subclinical AF detection and test the feasibility of a novel practice model with neurologists as ICM implanters. This unique pathway of clinical evaluation and delivery of cryptogenic stroke care, was briefly presented by the authors in the past, ^11, 17^ and the current report is the first published largest patient-series of ICM implants performed by non-cardiology service in the United States. The Nordic Atrial Fibrillation and Stroke (NOR-FIB) study^18^ conducted in 18 centers across Norway, Denmark and Sweden evaluated AF detection and burden in cryptogenic stroke and TIA using ICMs over a 12-month period. The NOR-FIB study also evaluated the feasibility of neurologists inserting ICMs in the stroke units and 91.5% of the ICMs in this study were implanted by a neurologist or stroke physician. In our study, VIN implanted ICMs only in the inpatient setting and performed 78-80% of all inpatient implants during the study period. Overall, 57.2% of all ICMs were performed by VIN and 42.8% by EP.

In acute care hospitals, particularly specialized stroke centers, collaboration between clinical specialties of neurology and cardiology in delivering stroke care is routinely observed. Conventionally, ICM implants in cryptogenic stroke for detection of AF is done by cardiologists, often with subspecialty expertise in electrophysiology. Neurologists play an integral role in the management of stroke and can potentially close the gaps in the evaluation and delivery of cryptogenic stroke care. Subspecialty training in vascular neurology further positions them well to improve patient selection and effectively determine ICM implant as reasonable and medically necessary. ICM implantation could potentially be considered as an extension of a cerebrovascular physicians’ clinical role. In our study, VIN served as additional physician-member in the care team with approved privileges to implant ICMs. This practice model may assist in avoiding possible delays in ICM implants, facilitate timely discharge of patients and potentially reduce the hospital length of stay. Additionally, having more implanters at a facility could close the gaps in access to stroke care at resource-limited hospitals, alleviate the burden on EP service particularly at busy stroke centers, and increase availability of service to ICM-eligible patients ready for discharge on the weekends and holidays. Both, the NOR-FIB and our study have clearly demonstrated the feasibility of neurologists as implanters of ICM. Furthermore, the observed procedural safety was high with a complication rate of 1.2% in the NOR-FIB and no complications in our study. ^18^

Using a collaborative care practice model, we demonstrated increased use of ICMs, particularly in the inpatient setting. This allowed for early start of uninterrupted long-term cardiac rhythm monitoring in patients. In our study, >75% of inpatient implants were done within 5 days and 95% within 10 days. The median time from index stroke to implant in the NOR-FIB study was comparable at 9 days. However, in the outpatient setting more than 50% of ICM implants in our study were performed ≥30 days after the diagnosis of cryptogenic stroke. This offers an opportunity to improve our outpatient ICM delivery process and simultaneously ensure external monitoring options as a bridge to ICM are offered. Delays in outpatient implantation could be related to prior authorization approvals for providing the service and/or availability of follow up appointments in cardiology EP clinic. It is important to note that traditional Medicare does not require, nor does it provide, prior authorization for subcutaneous long-term cardiac rhythm monitoring in eligible stroke patients.

Detection of AF leads to increased use of anticoagulation and prevention of recurrent stroke^7, 19^ and the use of ICMs increases detection of AF. ^19^ During the study period using a novel clinical practice model, both inpatient and total ICM implants increased by 141% and 130%, respectively. Simultaneously, the rates of AF detection also increased from 19.6% to 36.5%. AF detection in the subgroup of patients with inpatient ICMs implanted by VIN was 26% and 36.5% in the first 12-month and 24-month periods, respectively. These rates are comparable to the 28.6% AF detection in 12 months reported in the NOR-FIB study.

AF-related ischemic stroke is nearly twice as likely to be fatal when compared to non-AF stroke, and more likely to have severe residual deficits. ^20^ Increased efforts therefore are needed to detect AF and reduce the secondary stroke burden. Hospitals and stroke centers should evaluate their existing delivery models of cryptogenic stroke care and initiate efforts to close any gaps, particularly in detecting AF. Limitations of this study may largely arise from its observational design and use of data from a single center. Benefits of ICM in AF detection have been previously established, ^12–14^ and this study uses nonprobability sample of hospital registry data to successfully outline an innovative strategy that is unconventional and uncommon in cryptogenic stroke care using ICMs. Clinical care delivery was reimagined through collaboration and coordination between specialties, extending the role of vascular and endovascular/interventional neurology as implanters of ICMs, and using a closed-loop communication system to improve access to long-term cardiac rhythm monitoring and AF detection. Use of multidisciplinary, collaborative, coordinated and accessible practice models exemplify patient-centered care and are associated with improved outcomes. ^21^ The growing interest in the cerebrovascular and cardiology community to increase early detection of AF should therefore be combined with innovative pathways to increase access to high yield diagnostic long-term cardiac rhythm monitoring and improve cryptogenic stroke care and outcomes in patients.

## Data Availability

Data referred to in the manuscript is available for review.

## Abbreviations and Acronyms

AF: Atrial fibrillation
ICM: Insertable Cardiac Monitor
EP: Electrophysiologist
VIN: Vascular and Interventional Neurologist

## Acknowledgments

None

## Sources of Funding

None

## Disclosures

None

## Notes

### Competing Interest Statement

Corresponding author is a consultant to Medtronic, Inc.

### Clinical Trial

N/A

### Funding Statement

Unfunded

### Author Declarations

Study is reviewed and approved by the Institutional Review Board of the Thomas Jefferson University.

